# High co-circulation of influenza and SARS-CoV-2

**DOI:** 10.1101/2022.09.13.22279740

**Authors:** John T Kubale, Aaron M Frutos, Angel Balmaseda, Saira Saborio, Sergio Ojeda, Carlos Barilla, Nery Sanchez, Abigail Shotwell, Alyssa Meyers, Roger Lopez, Miguel Plazaola, Guillermina Kuan, Aubree Gordon

**Author notes:** **Corresponding author:** Aubree Gordon: School of Public Health, University of Michigan, Ann Arbor, MI 48109. **Alternate corresponding author:** John Kubale: ICPSR, University of Michigan, Ann Arbor, MI 48109.

## Abstract

In the first two years of the COVID-19 pandemic, influenza transmission decreased substantially worldwide meaning that health systems were not faced with simultaneous respiratory epidemics. In 2022, however, substantial influenza transmission returned to Nicaragua where it co-circulated with SARS-CoV-2 causing substantial disease burden.

---

Early in the COVID-19 pandemic, influenza circulation collapsed globally, including in Nicaragua where only 5 cases of influenza (80% influenza B) were detected in 2021.[1, 2] While concerns about the possibility of influenza and SARS-CoV-2 co-circulation were raised in the lead up to the typical Northern hemisphere influenza season in late 2020 and 2021, they did not materialize. Recently, however, substantial influenza transmission has returned to the Southern Hemisphere and tropical settings including Nicaragua suggesting this reprieve is likely over.[3] While vaccine coverage during the early phase of the pandemic was higher than pre-pandemic levels, it decreased last season (2021/22). [2, 4, 5] Given the resurgence of influenza in 2022, this represents a worrying trend as the typical Northern Hemisphere influenza season approaches. Here we describe substantial influenza and SARS-CoV-2 co-circulation within a prospective, community-based household study in Managua, Nicaragua and consider its implications for the looming fall/winter season in the Northern Hemisphere.

## Methods

The Household Influenza Cohort Study is an ongoing, community-based prospective cohort study in Managua, Nicaragua.[6] This study was approved by the institutional review boards at the University of Michigan and the Nicaraguan Ministry of Health. Informed consent and parental approval (for minors) was obtained for all participants and assent was obtained from all children aged ≥6 years. Participants presented to the study clinic upon the development of an acute illness and respiratory samples were collected from those meeting the testing criteria (fever/feverishness, conjunctivitis, rash, or loss of taste or smell). Respiratory samples were tested for influenza (using CDC protocols) and SARS-CoV-2 by real time RT-PCR.[7] Samples were also collected from household members, regardless of symptoms, following the positive test (for influenza or SARS-CoV-2) of another household member.[6]

### Clinical definitions

Illness severity was classified using symptom diaries and data from clinic visits.[7] Specifically, illnesses involving hospitalization, difficulty or rapid breathing, crepitus, chest wall indrawing, rhonchi, wheezing, and overall poor condition were classified as moderate/severe, while those with no symptoms or other symptom presentations were classified as mild/asymptomatic. Those requiring transfer to the hospital within 28 days of illness onset were classified as hospitalized. To assess whether the number of symptomatic influenza/SARS-CoV-2 co-infections we observed differed from the number we would expect (if circulation was independent) we pooled samples from the Household Influenza Cohort with those from the Nicaraguan Pediatric Influenza Cohort Study who met the same testing criteria for symptomatic illness. Samples positive for both influenza and SARS-CoV-2 (via real time RT-PCR) were considered co-infections.

### Statistical analysis

Incidence rates were calculated using a Poisson distribution[8] while the observed and expected number of co-infections were compared using the chi-squared test. The attack rates for influenza A/H3N2 and SARS-CoV-2 were calculated among those participants who were enrolled during the entire study period (number of cases of each pathogen/total number of participants). To assess what these attack rates would look like in the US we standardized estimates using the 2022 World Population Prospects from the United Nations. All statistical analyses were completed using R version 4.2.1.

## Results

We examined influenza SARS-CoV-2 infections and co-infections among 2117 participants (62.5% female) aged 0-89 years from January 1-July 20, 2022. Overall, there were 433 influenza A/H3N2 infections (incidence rate of 37.6 per 100 person-years; 95% confidence interval [CI]: 34.1, 41.3), and 296 SARS-CoV-2 infections (26.0 per 100 person-years; 95% CI: 23.1, 29.1). Rates of influenza peaked among the youngest participants (aged <5 years) and steadily decreased thereafter. Rates of SARS-CoV-2 by age displayed a slight V-shaped trend (Figure 1). We observed no meaningful difference in incidence by sex (Supplemental table 1). Looking at detections by household, 174 (40.1%) households experienced influenza A/H3N2, 105 (28.2%) had SARS-CoV-2, and 38 (10.7%) had both.

**Figure 1:**
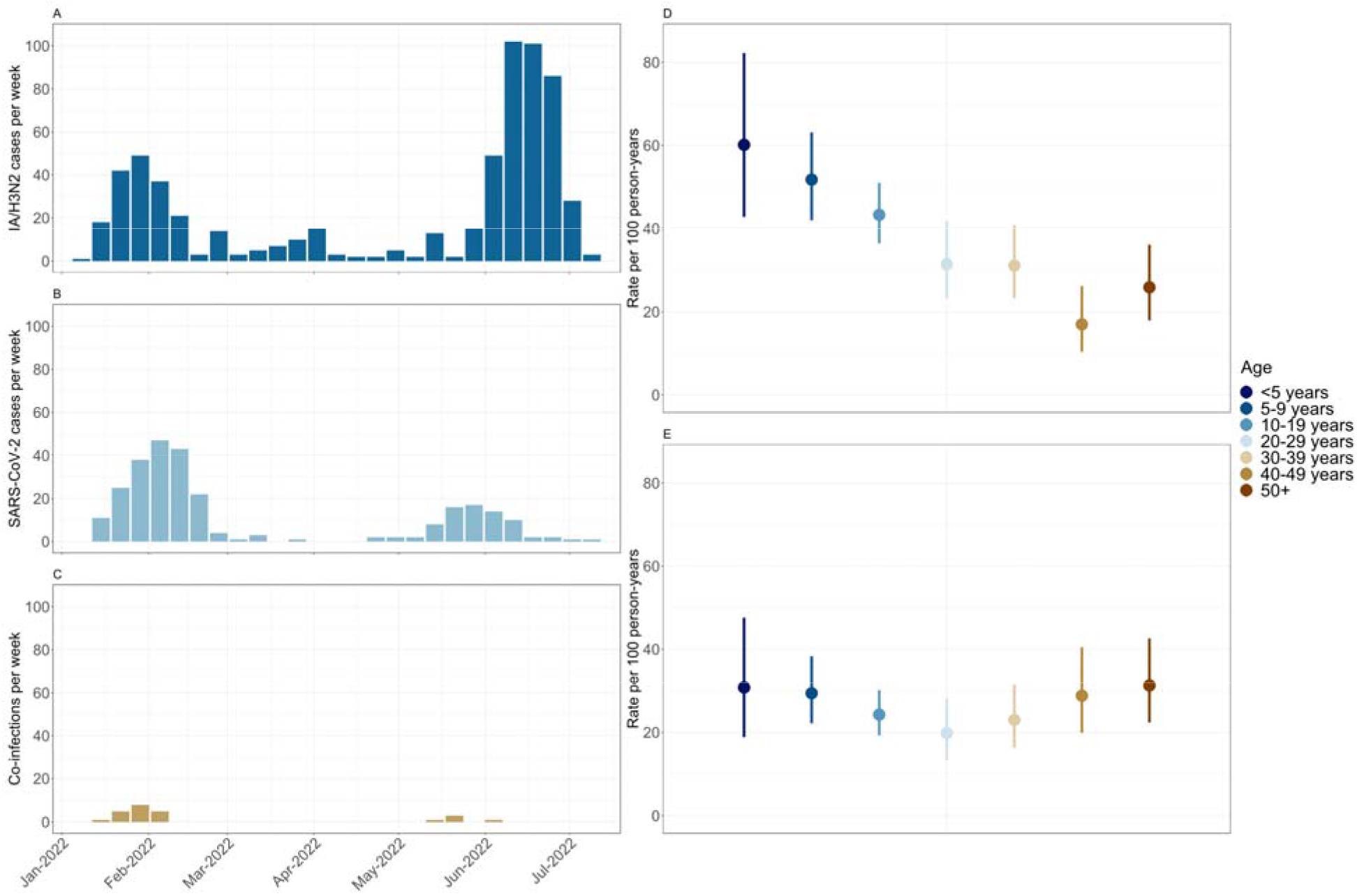
Influenza and SARS-CoV-2 in the Cohort. Panels A-C show the number of cases per week for influenza A/H3N2, SARS-CoV-2, and H3N2/SARS-CoV-2 co-infections respectively. Panels D and E show the incidence rate (per 100 person years) by age for influenza A/H3N2 and SARS-CoV-2 respectively.

### Clinical presentation and severity

In total, 3 participants required hospitalization (2 with SARS-CoV-2, 1 with influenza A/H3N2 infections). No co-infected participants required hospitalization. A greater proportion of SARS-CoV-2 cases were classified as moderate/severe compared to influenza A/H3N2 (9.6% vs 4.2%, p=0.004), despite the study population having high levels of hybrid immunity. However, no difference was observed among children (3.4% vs 5.5%, p = 0.4, Supplemental Table 3) nor when hospitalizations were compared (0.7% vs 0.2%, p=1.0, Supplemental Table 3). While the most frequent symptom combinations were similar across infection types (fever and upper respiratory symptoms [Supplemental Figure 1]), a greater proportion of SARS-CoV-2 infections presented with cough, myalgia, and arthralgia compared to influenza (Supplemental Table 2). However, a greater proportion of co-infected participants did have fever when compared to those with SARS-CoV-2 single infections (p=0.03).

### Dual burden

Influenza A/H3N2 and SARS-CoV-2 co-circulated for 22/29 (75.9%) of the study weeks. The influenza attack rate was 20.1% (95% CI: 18.4, 21.8) while the attack rate of SARS-CoV-2 was 13.6% (95% CI: 12.2, 15.1) (Supplemental Table 4). When standardized to the age distribution of the United States which is older than Nicaragua and our cohort, we found similarly high attack rates, specifically 17.2% (95% CI: 14.0, 20.4) for influenza and 14.3% (95% CI: 12.7, 16.0) for SARS-CoV-2. In children aged 2-14 years, the attack rate of influenza was 26.8% (95% CI: 23.7, 29.9) compared to an attack rate of 15.3% (95% CI: 12.7, 17.8) for SARS-CoV-2. Indeed, when compared to prior influenza years in the cohort (overall 14.5 per 100 person years; range 8.0 to 21.6)[9] the 2022 incidence rate to date, assuming no additional circulation, is substantially higher at 28.6 (95% CI: 25.0, 32.5) per 100 person years. We observed approximately the expected number of symptomatic influenza/SARS-CoV-2 co-infections (p=0.39 Supplemental Table 5).

## Discussion

Here we observed substantial simultaneous burden of influenza A/H3N2 and SARS-CoV-2 within a prospective, community-based household cohort in Managua, Nicaragua. Influenza and SARS-CoV-2 co-circulated for most of the study period and the number of co-infections was near what we would expect if the distribution of the pathogens were independent. This suggests limited viral interference, and that the primary danger of co-circulation is high rates of single infections occurring concurrently. In fact, the estimated attack rate for influenza in children aged 2 to 14 years (26.8%) was higher than that seen in this population during the 2009 H1N1 pandemic [10], and this was on top of a SARS-CoV-2 attack rate of 13.6%. Taken together, this represents a substantial overall burden on the health system. When standardized to the age distribution of the United States the influenza attack rate is slightly lower and SARS-CoV-2 is slightly higher, though the differences were not significant. However, it is also important to consider that SARS-CoV-2 seroprevalence also plays an important role. In Nicaragua, the majority of the population has previously been infected with SARS-CoV-2, and many have also been vaccinated.[11] Given the older age distribution in the US we anticipate that similar levels of co-circulation may in fact lead to greater rates of illness and severe disease.

The high attack rates in children are also concerning as they suggest substantial morbidity and further school disruptions. Further, pediatric influenza vaccination coverage has steadily decreased since the start of the pandemic, even when adult vaccination coverage remained high. Additionally, though vaccines against SARS-CoV-2 have been approved for children in the US, vaccination coverage remains quite low among those aged <12 years. In fact, only 38% of 5–11-year-olds and 7% of children 6 months—4 years have received at least one COVID-19 vaccine dose.[12]

This study has several strengths. First, as a longitudinal, community-based study we were able to calculate incidence rates of both SARS-CoV-2 and influenza in the population. Second, the study design involved testing asymptomatic participants following household activation which improves the accuracy of these incidence measures by better capturing subclinical infections. Finally, studies have explored the burden and transmission of influenza in this community for over fifteen years providing important context for these new estimates.

This study does have some limitations. While we failed to detect a difference in the number of observed and expected co-infections the relatively small number (n=48) and follow up period <1 year precludes us from ruling out the possibility of viral interference, or a synergistic effect for that matter. Additionally, we were only able to assess the number of symptomatic co-infections, so this likely represents an underestimate of the total co-infection burden (asymptomatic and symptomatic). While we recognize the importance of accounting for asymptomatic co-infections in assessing transmission, we contend that symptomatic co-infections are a reasonable means of assessing relative burden when combined with more comprehensive measures of single-infections (i.e. that capture asymptomatic infections). Finally, generalizing these findings to other populations should be done with appropriate consideration of differences in population-level immunity to both SARS-CoV-2 and influenza and the means through which the immunity was obtained (i.e. infection and/or vaccination).

In this study we describe substantial concurrent circulation of influenza and SARS-CoV-2 within a prospective, community-based cohort. These findings suggest that increased susceptibility to influenza after low-circulation places populations at significant risk of having dual epidemics of influenza and SARS-CoV-2. That this is likely to be worse in populations with lower prior SARS-CoV-2 infection rates, further highlights that vaccination against both SARS-CoV-2 and influenza is imperative this coming season.

## Data Availability

Individual-level data may be shared with outside investigators following University of Michigan IRB approval. Please contact Aubree Gordon to arrange for data access.

## Acknowledgements

The authors would like to thank the study participants and their families along with the study staff at HCSFV and the Centro Nacional de Diagnóstico y Referencia.

