## Supplemental Materials for "High co-circulation of influenza and SARS-CoV-2"

**Table of Contents:**

Page 3) Supplemental Figure 1: Most common symptom combinations by infection type

Page 4) Supplemental Table 1: Influenza and SARS-CoV-2 incidence rates by sex

Page 5) Supplemental Table 2: Symptom presentation and treatment within 28 days of infection detection

Page 6) Supplemental Table 3: Severity of influenza A/H3N2 and SARS-CoV-2 infections

Page 7) Supplemental Table 4: Attack rates of influenza A/H3N2 and SARS-CoV-2

Page 8) Supplemental Table 5: Observed and expected number of Influenza A/H3N2 and COVID-19 co-infections

**Supplemental Figure 1: Most common symptom combinations by infection type**

Panel A shows the most common symptom combinations observed in influenza A/H3N2 infections, Panel B shows the most common symptom combinations observed in SARS-CoV-2 infections, Panel C shows the most common symptom combinations observed in H3N2/SARS-CoV-2 co-infections.

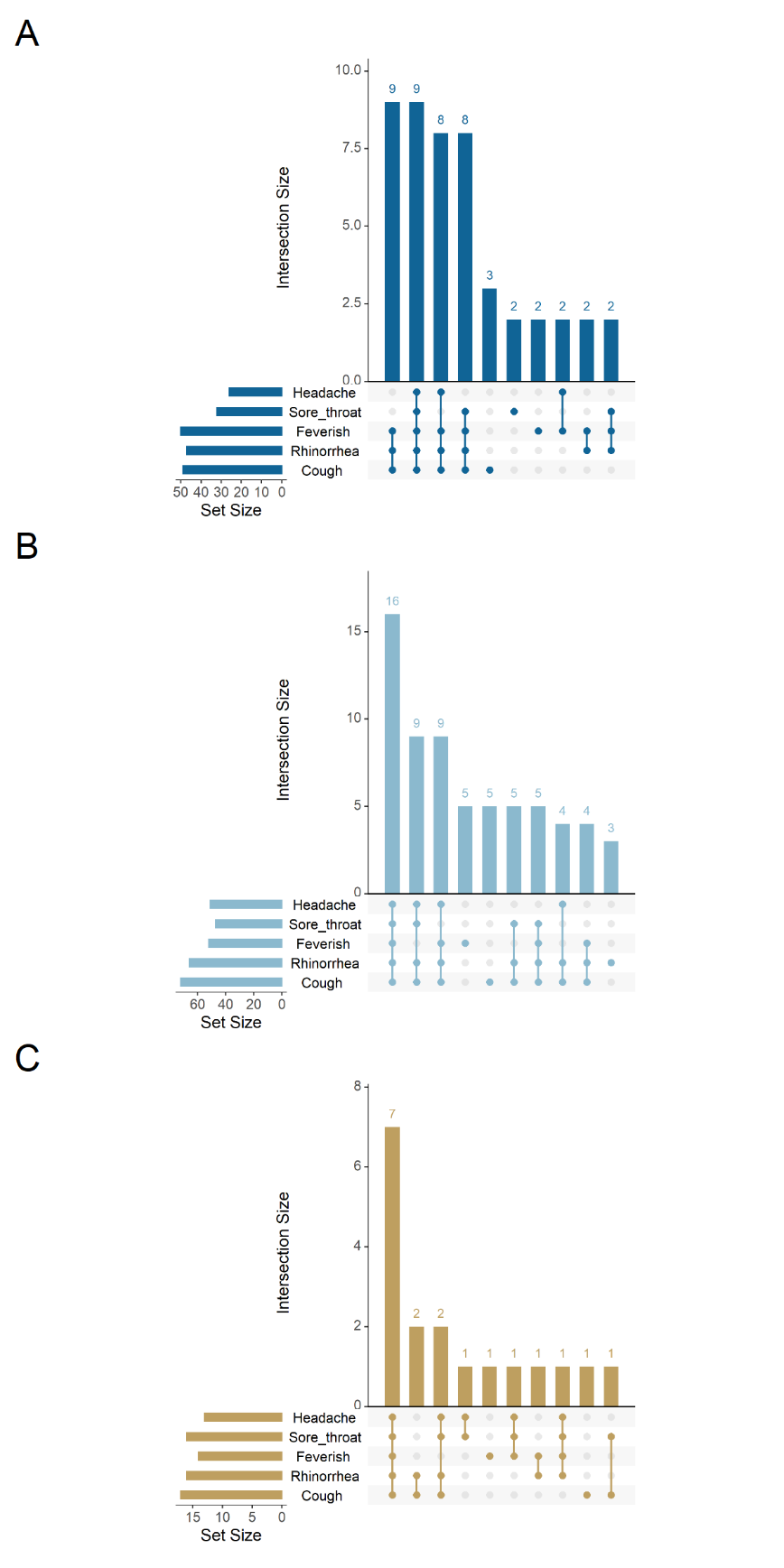

**Supplemental Table 1: Influenza and SARS-CoV-2 incidence rates by sex**

|  | Cases | Person years | Incidence Rate (95% CI) per 100 person years |
| --- | --- | --- | --- |
| **Influenza A/H3N2** |  |  |  |
| Male | 175 | 451 | 38.82 (33.28, 45.01) |
| Female | 258 | 701 | 36.82 (32.47, 41.60) |
| **SARS-CoV-2** |  |  |  |
| Male | 120 | 446 | 26.89 (22.29 32.15) |
| Female | 176 | 694 | 25.35 (21.75 29.39) |

**Supplemental Table 2: Symptom presentation and treatment within 28 days of infection detection**

|  | | | | | |
| --- | --- | --- | --- | --- | --- |
| Variable | Overall | Influenza A | SARS-CoV-2 | Co-infection | p-value* |
| No. | 705 | 409 | 272 | 24 |  |
| Feverish | 214 (30.4) | 115 (28.1) | 83 (30.5) | 16 (66.7) | 0.0007 |
| Measured fever | 103 (14.6) | 57 (13.9) | 39 (14.3) | 7 (29.2) | 0.0734 |
| Cough | 274 (38.9) | 118 (28.9) | 137 (50.4) | 19 (79.2) | 0.0126 |
| Rhinorrhea | 255 (36.2) | 117 (28.6) | 120 (44.1) | 18 (75.0) | 0.0071 |
| Sore throat | 161 (22.8) | 63 (15.4) | 81 (29.8) | 17 (70.8) | 0.0001 |
| Congestion | 128 (18.2) | 46 (11.2) | 70 (25.7) | 12 (50.0) | 0.0210 |
| Headache | 137 (19.4) | 41 (10.0) | 82 (30.1) | 14 (58.3) | 0.0093 |
| Loss of appetite | 62 (8.8) | 15 (3.7) | 39 (14.3) | 8 (33.3) | 0.0350 |
| Myalgia | 66 (9.4) | 19 (4.6) | 40 (14.7) | 7 (29.2) | 0.0785 |
| Arthralgia | 50 (7.1) | 10 (2.4) | 33 (12.1) | 7 (29.2) | 0.0289 |
| Rapid breathing | 5 (0.7) | 1 (0.2) | 3 (1.1) | 1 (4.2) | 0.2883 |
| Diarrhea | 16 (2.3) | 6 (1.5) | 9 (3.3) | 1 (4.2) | 0.5765 |
| Hospitalized | 3 (0.4) | 1 (0.2) | 2 (0.7) | 0 (0.0) | 1.0000 |
| Antibiotics used | 30 (4.3) | 12 (2.9) | 15 (5.5) | 3 (12.5) | 0.1700 |
| *p-value is from a chi-square or Fisher's exact test for SARS-CoV-2 and co-infections | | | | | |

**Supplemental Table 3: Severity of influenza A/H3N2 and SARS-CoV-2 infections**

| **Overall** | | | |
| --- | --- | --- | --- |
|  | Moderate/Severe | Asymptomatic/Mild | Total |
| Influenza A/H3N2 | 17 | 392 | 409 |
| SARS-CoV-2 | 26 | 246 | 272 |
| Total | 43 | 638 |  |
| P = 0.0039 |  |  |  |
| **<15 years** | | | |
|  | Moderate/Severe | Asymptomatic/Mild | Total |
| Influenza A/H3N2 | 12 | 205 | 217 |
| SARS-CoV-2 | 4 | 114 | 118 |
| Total | 16 | 319 |  |
| P=0.4069 | | | |

**Supplemental Table 4: Attack rates of influenza A/H3N2 and SARS-CoV-2**

| **This study** | | |
| --- | --- | --- |
|  | Attack Rate (aged 0-89 years) | Attack Rate (aged 2-14 years) |
| Influenza A/H3N2 | 20.1% (18.4, 21.8) | 26.8% (23.7, 29.9) |
| SARS-CoV-2 | 13.6% (12.2, 15.1) | 15.3% (12.7, 17.8) |
| **2009 H1N1 pandemic^1^** | | |
|  | Attack Rate (aged 0-89 years) | Attack Rate (aged 2-14 years) |
| Influenza A/H1N1 | -- | 20.1% (18.8, 21.4) |

**Supplemental Table 5: Observed and expected number of Influenza A/H3N2 and COVID-19 co-infections**

| **Observed** | | | |
| --- | --- | --- | --- |
|  | Influenza A/H3N2 | No Influenza A/H3N2 | Total |
| COVID-19 | 48 | 199 | 247 |
| No COVID-19 | 372 | 1311 | 1683 |
| Total | 420 | 1510 |  |
| **Expected** | | | |
|  | Influenza A/H3N2 | No Influenza A/H3N2 | Total |
| COVID-19 | 53.8 | 193.2 | 247 |
| No COVID-19 | 366.2 | 1316.8 | 1683 |
| Total | 420 | 1510 |  |
